# Mechanistic Transmission Modeling of COVID-19 on the Diamond Princess Cruise Ship Demonstrates the Importance of Aerosol Transmission

**DOI:** 10.1101/2020.07.13.20153049

**Authors:** Parham Azimi, Zahra Keshavarz, Jose Guillermo Cedeno Laurent, Brent R. Stephens, Joseph G. Allen

## Abstract

**Background:** The current prevailing position is that coronavirus disease 2019 (COVID-19) is transmitted primarily through large respiratory droplets within close proximity (i.e., 1-2 m) of infected individuals. However, quantitative information on the relative importance of specific transmission pathways of the severe acute respiratory syndrome coronavirus 2 (SARS-CoV-2) (i.e., droplets, aerosols, and fomites across short- and long-range distances) remains limited.

**Methods:** To evaluate the relative importance of multiple transmission routes for SARS-CoV-2, we leveraged detailed information available from the Diamond Princess Cruise Ship outbreak that occurred in early 2020. We developed a framework that combines stochastic Markov chain and negative exponential dose-response modeling with available empirical data on mechanisms of SARS-CoV-2 dynamics and human behaviors, which informs a modified version of the Reed-Frost epidemic model to predict daily and cumulative daily case counts on the ship. We modeled 21,600 scenarios to generate a matrix of solutions across a full range of assumptions for eight unknown or uncertain epidemic and mechanistic transmission factors, including the magnitude of droplet and aerosol emissions from infected individuals, the infectious dose for deposition of droplets and aerosols to the upper and lower respiratory tracts, and others.

**Findings:** A total of 132 model iterations met acceptability criteria (R^2^ > 0.95 for modeled vs. reported cumulative daily cases and R^2^ > 0 for daily cases). Analyzing only these successful model iterations yields insights into the likely values for uncertain parameters and quantifies the likely contributions of each defined mode of transmission. Mean estimates of the contributions of short-range, long-range, and fomite transmission modes to infected cases aboard the ship across the entire simulation time period were 35%, 35%, and 30%, respectively. Mean estimates of the contributions of large respiratory droplets and small respiratory aerosols were 41% and 59%. Short-range transmission was the dominant mode after passenger quarantine began, albeit due primarily to aerosol transmission, not droplets.

**Interpretation:** Our results demonstrate that aerosol inhalation was likely the dominant contributor to COVID-19 transmission among passengers aboard the Diamond Princess Cruise Ship. Moreover, close-range and long-range transmission likely contributed similarly to disease progression aboard the ship, with fomite transmission playing a smaller role. The passenger quarantine also affected the importance of each mode, demonstrating the impacts of the interventions. Although cruise ships represent unique built environments with high ventilation rates and no air recirculation, these findings underscore the importance of implementing public health measures that target the control of inhalation of aerosols in addition to ongoing measures targeting control of large droplet and fomite transmission, not only aboard cruise ships but in other indoor environments as well.

**Funding:** Funding information is not available.

## Introduction

Information on the relative importance of specific transmission pathways of the severe acute respiratory syndrome coronavirus 2 (SARS-CoV-2) remains limited.^1^ The World Health Organization’s (WHO) current position is that the COVID-19 virus is transmitted primarily through respiratory droplets and contact routes, while airborne transmission of the COVID-19 virus is likely not a major route of transmission other than in settings in which aerosol generating procedures are occurring.^2^ Similarly, the U.S. Centers for Disease Control and Prevention (CDC) recently updated their position to “COVID-19 is thought to spread mainly through close contact from person-to-person,” which CDC defines as within about 1.8 m (6 feet), and that fomite transmission and inhalation of respiratory droplets are likely not the main ways that the virus spreads.^3^

Conversely, numerous researchers^4–12^ and professional societies (e.g., ASHRAE^13^) have raised concerns that longer-range airborne transmission is likely occurring from both symptomatic and asymptomatic (or pre-symptomatic) individuals through a combination of larger respiratory droplets that are carried further than 1-2 m via airflow patterns and smaller inhalable aerosols (i.e., ‘droplet nuclei’) that can easily transport over longer distances. These concerns arise from a growing understanding of human respiratory emissions,^14,15^ known transmission pathways of other respiratory viruses,^16^ recent empirical evidence detecting SARS-CoV-2 in aerosol and surface samples in healthcare settings,^17–20^ and recent case studies demonstrating the likely importance of longer-range aerosol transmission in some settings.^21,22^ Understanding the importance of each transmission pathway for COVID-19 is critical to informing public health guidelines for effectively managing the spread of the disease.

In the absence of empirical studies using controlled exposures to elucidate transmission pathways,^23^ mathematical modeling approaches can offer insights into the likely importance of the different modes of disease transmission among human populations,^24–28^ provided that sufficiently accurate inputs are available. To help fill these knowledge gaps, this work uses a mechanistic modeling approach to investigate the relative importance of multiple transmission routes of SARS-CoV-2 among individuals aboard the Diamond Princess Cruise Ship, which experienced a major outbreak of COVID-19 in early 2020.

## Methods

The Diamond Princess Cruise Ship presents a unique built environment case study, with a known number of passengers, crewmembers, and COVID-19 cases over time, discovered through high rates of testing, and a relatively high degree of knowledge of several important human and built environment factors. The Diamond Princess experienced a major outbreak of COVID-19 in early 2020, with 712 of 3711 passengers and crew members on board becoming infected (19% of the community)^29^ and at least 57 other passengers who tested positive in the days after they left the ship and returned to their home countries.^30^ As reported, the COVID-19 outbreak was traced to a single passenger from Hong Kong who boarded the ship in Yokohama on January 20 and then disembarked in Hong Kong on January 25. He had symptoms including coughing before boarding and was diagnosed with COVID-19 on February 1 in Hong Kong. The first 10 cases were confirmed on February 4 after the ship arrived in the Yokohama port. Laboratory-confirmed cases of COVID-19 led to the quarantine of passengers aboard the Diamond Princess for 14 days beginning on February 5 at 7 am, with all passengers required to remain in their cabins essentially all of the time. As of February 5, there were a total of 3711 individuals onboard the Diamond Princess, with 2666 passengers and 1045 crew members.^31^

To estimate the likely contributions of specific infection transmission modes to the number of COVID-19 cases among individuals aboard the Diamond Princess Cruise Ship, a combination of epidemic, mechanistic transmission, and dose-response models was adopted. Full model details are described in the SI. Briefly, we utilize a stochastic Markov chain process to stochastically trace close- and long-range transmission by contact with large respiratory droplets, inhalation of smaller aerosols, and fomite contact under a wide range of possible scenarios constructed from combinations of unknown or uncertain input parameters. The Markov chain model informs a dose-response model, which in turn informs an epidemic model to generate estimates of daily and cumulative daily case counts aboard the ship from January 20 (when there was only one index case aboard the ship) to February 24 (when all passengers disembarked). We analyze only those model scenarios that achieved an acceptable agreement between predicted and reported case numbers for daily cumulative cases (defined as R^2^ > 0.95) and daily cases (defined as non-negative R^2^) to infer likely values of the unknown or uncertain model parameters and to quantify the contribution of the various modes of transmission in the most successful model scenarios.

### Markov Chain Model

The mechanistic transmission model uses a Markov chain process to estimate the number of SARS-CoV-2 copies present in numerous physical states, as well as the probability of transmission of SARS-CoV-2 between each defined state, aboard the ship over time (SI Section 1). We considered 12 *states* for the Markov chain process, including indoor air and surfaces in cabins and public areas, hands (palms) of individuals, upper and lower respiratory tracts (i.e., URT and LRT) of individuals, heating, ventilation, and air-conditioning (HVAC) systems, and inactivation of viable virus (Figure S1). We generated a new Markov chain matrix (MCM) for each day in the simulation period to model mechanistic transmission and infection probability based on a number of assumptions for built environment parameters, crew and passengers’ interactions, adopted infection control strategies, and the number of infectors and susceptible individuals estimated from application of the transmission risk model to the previous days.

The modeling framework incorporates available empirical data on key mechanisms of SARS-CoV-2 dynamics culled from recent literature, including (i) viral RNA emission rates in large droplets (> 10 µm) and inhalable aerosols (< 10 µm) from infected individuals, which were back-calculated from recent reports of air and surface sampling in healthcare settings and were assumed to be the same ratio for all infected individuals, (ii) viability loss in air and on surfaces reported in controlled studies, and (iii) estimates of aerosol deposition rates to surfaces based on typical assumptions for aerosol dynamics.

The framework also leverages estimates and assumptions for several human and built environment transmission factors, culled from prior literature where possible, including average rates of face- and surface-touching, inhalation rates, the shape and size of close-contact zones, time spent in various environments (e.g., public areas and cabins), floor areas and volumes of cabins and public areas, the probability of uninfected individuals within close proximity of an infected individual, and the impact of infection control strategies that were implemented during the quarantine period (e.g., mask wearing, hand washing, and surface disinfection). Detailed descriptions of all model inputs are provided in the SI, including Section 1 (for relatively certain parameters) and Section 3 (for relatively unknown or uncertain parameters).

### Dose-Response Model

To estimate the infection probability of SARS-CoV-2 viruses deposited to different body sites of susceptible individuals, we used a negative exponential dose-response model, which implies that a single particle can start an infection and all single particles are independent of each other. The probability of infection for one susceptible individual (*P*_*infection*_) in the cruise ship was calculated using Equation 1:

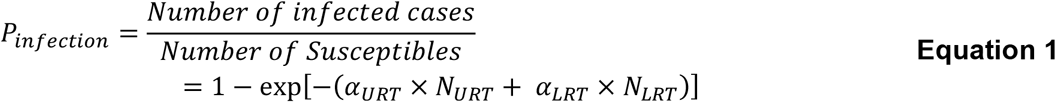

Where:

*N*_*URT*_ and *N*_*LRT*_: Number of viable SARS-CoV-2 RNA copies in upper and lower respiratory tracts of one susceptible individual, and

*α*_*URT*_ and *α*_*LRT*_: Infectivity of SARS-CoV-2 for upper and lower respiratory tracts.

The 50% infectious dose (*ID*_50_), or the number of viruses necessary to infect a susceptible individual in 50% of a sample population, of SARS-CoV-2 for upper and lower respiratory tracts can be estimated from Equation 2:^32,33^

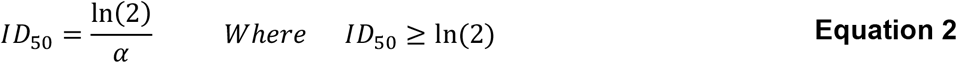

Estimates of ID_50_ and infectivity for upper and lower respiratory tracts (URT and LRT) play a critical role in understanding the transmission of airborne infectious diseases. However, we are not aware of any clinical studies to date that report these values for SARS-CoV-2 in humans or animals. Moreover, the proportions of SARS-CoV-2 depositing in the LRT and URT of a susceptible individual when they inhale infectious aerosols are not yet characterized. Therefore, we tested three logarithmically spaced assumptions for the ratio of the effective ID_50_ for SARS-CoV-2 for aerosol inhalation (assuming deposition in the LRT) and fomite and droplet deposition (assuming deposition in the URT) (i.e., ID_50_ URT:LRT = 1:1, 10:1, and 100:1). We rely on our model approach to back-calculate effective ID_50_ values (using a basis of RNA copies) by analyzing successful model results, as described in the SI (Section 1.3). This approach allows us to test scenarios with this uncertain parameters without knowing (or needing to know) the actual magnitude of ID_50_, which can then be used to infer the likely magnitude of this ratio based on successful model outcomes.

### Transmission Mode Contribution to Infection

In addition to estimating the number of infected cases with the model framework, we also estimated the contribution of multiple infection transmission modes to the estimated number of infected cases in both cabins and public areas, including (i) direct deposition of respiratory droplets (within close range only), (ii) fomite transmission, and (ii) inhalation of aerosols (with both close- and long-range transmission traced separately) (Equation 3):

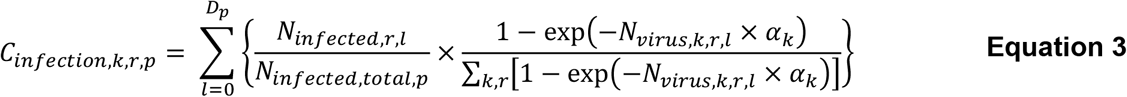

Where:

*k*: Four considered scenarios for infection transmission modes, including direct droplet deposition, fomite, long-range aerosol inhalation, and short-range aerosol inhalation,

*r*: Two considered micro-environments in the cruise ship including cabins and public areas,

*p*: Three considered simulation periods including during the entire outbreak duration, before the passenger quarantine began, and after the passenger quarantine began,

*C*_*infection,k,r,p*_: Infection contribution associated with transmission mode *k* in microenvironment *r* in simulation period *p*,

*D*_*p*_: Number of simulation days in the simulation period *p* (i.e., 36, 16, and 20 for the entire outbreak duration before all passengers disembarked, before the passenger quarantine began, and after the passenger quarantine began, respectively),

*N*_*infection r,l*_: Number of infected cases in microenvironment *r* on day of the simulation period,

*N*_*infection,total,p*_: Total number of infected cases in the cruise ship during the simulation period *p*,

*N*_*virus,k,r,l*_ : Number of SARS-CoV-2 RNA copies that reached the relevant respiratory tract region (i.e., LRT for inhalation and URT for direct deposition and fomite) via transmission mode *k* in microenvironment *r* on day of the simulation period, and

*α*_*k*_: Infectivity of SARS-CoV-2 for the target respiratory tract (i.e., LRT for inhalation and URT for direct deposition and fomite)

This approach allows for summarizing estimates of infection contributions by transmission mode, contact range, micro-environment (i.e., public areas or passenger cabins), and/or simulation period independently, as needed.

Short-range transmission occurs by direct deposition of respiratory droplets and inhalation of aerosols only when susceptible individuals were within a defined close-range contact area of infected individuals. The close-range contact area was defined assuming a conical area in front of an infector with the head angle of 60° and length of 3 meters (described in detail in the SI, Section 1.2.2)^34,35^. The projected surface area of the cone on the floor was ∼4.7 m^2^, which is equivalent to a surface area of a circle around the infector with a radius of ∼1.2 m. The probability that a susceptible individual was present within the close-contact cone was estimated based on the proportion of the zone surface area to the projected surface area of the cone on the floor (SI Section 1.2.2).

Long-range inhalation transmission occurs via inhalation of aerosols when susceptible individuals were outside the close-contact area. Fomite transmission occurs when susceptible individuals came in contact with contaminated surfaces, which could be contaminated by infected individuals through direct touching, direct deposition of respiratory droplets, and/or deposition of respiratory aerosols at any time point and location in the model framework.

### Combining the Transmission Risk Model with a Developed Epidemic Model

The mechanistic infection transmission model was combined with a modified version of the Reed-Frost epidemic model to simulate the transmission of COVID-19 aboard the ship. We assumed that (i) the infection is spread from infected individuals to others by four main transmission pathways (long-range inhalation, short-range inhalation, direct deposition within close-range, and fomite), (ii) a portion of susceptible individuals in the group will develop the infection and will be infectious to others (the portion of ‘*susceptibles’* who will develop the infection is estimated by the transmission risk model), (iii) the probability of coming into adequate contact with any other specified individual in the group within one time interval depends on the interaction behavior of the individual and is estimated using the Markov chain method, (iv) the susceptible individuals in the cruise ship were isolated from others outside the cruise ship, and (v) these conditions remain constant during one whole day of the outbreak.

To estimate the spread of the disease, we estimated the number of infected cases among susceptible individuals, some of whom were cabinmates with infected individuals and some were not, at the end of each simulation day using the transmission risk model. The infected cases were assumed to develop infection and become *‘infectors’* after the latent period, which was estimated by reducing the assumed effective sub-clinical infectious period (i.e., the time span between the onset of the infectious period and the appearance of clinical signs of disease) from the effective incubation period (i.e., the time span between infection and detection among infected cases). The number of cabins with at least one infected individual (i.e., ‘infected cabins’) was calculated at the end of each simulation day by assuming the number of newly infected cabins is equal to the number of newly infected cases who were not in one of the previously infected cabins at the beginning of the simulation day. The numbers of susceptible individuals who were not cabinmates with an infector (*N*_*susceptibles*−*common*_) and susceptible individuals inside the infected cabins (*N*_*susceptibles*−*cabin*_) at the beginning of each simulation day (*d*) were estimated using the Equations 4-5 (except for the first period of infection transmission):

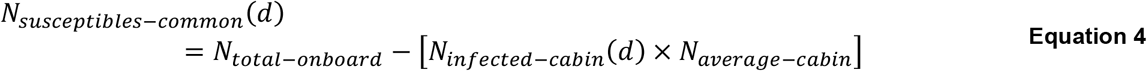

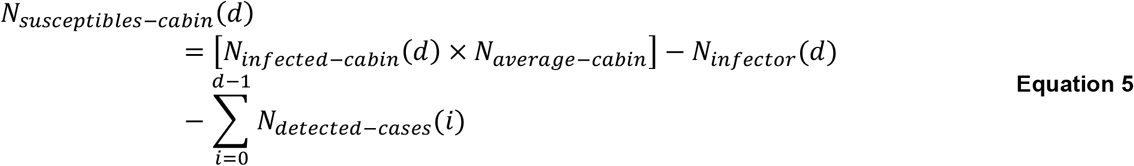

Where:

*N*_*total* −*onboard*_ : Total number of passengers and crew onboard (constant during the outbreak),

*N*_*infection* − *cabin*_: Estimated number of infected cabins at the beginning of each day,

*N*_*average* − *cabin*_: Average number of individuals in one cabin,

*N*_*infecter*_: Number of infectors, and

∑ *N*_*dedected* −*cases*_: Cumulative number of detected infected cases or disembarked individuals from the cruise ship.

We assumed the infected cases could spread infectious particles only one day after the incubation period, when their clinical symptoms began. We divided the transmission patterns into four periods, each of which having different epidemic characteristics, as described in the SI (Section 1.1). Several checkpoint conditions were introduced to the epidemic model to ensure reasonable bounds (SI Section 2.3).

### Analysis

The model framework requires numerous assumptions or estimates for unknown or uncertain input parameters, which were culled from existing literature where possible and otherwise estimated or assumed using known information about the Diamond Princess Cruise Ship. Because there is high uncertainty around several critical model parameters, we utilized a scenario modeling approach in which values for unknown or uncertain epidemic and transmission modeling parameters were varied over a wide range of possibilities to generate a matrix of possible solutions. A total of 21,600 scenarios were modeled across a range of estimates or assumptions for eight critical unknown or uncertain input parameters (Table 1). Estimates and assumptions for these parameters are described in detail in the SI (Section 3). We ran the model with each possible combination of the eight unknown or uncertain input parameters shown in Table 1 (10×5×6×3×3×2×2×2=21,600) in order to search a wide range of possible parameter values and combinations of parameter values.

**Table 1.** Summary of the ranges of 8 unknown or uncertain critical model input parameters that defined each model iteration

| Model Inputs | Epidemiological Factors |  |  | Mechanistic Transmission Factors |  |  |  |  |
| --- | --- | --- | --- | --- | --- | --- | --- | --- |
|  | Effective incubation period | Effective sub-clinical infectious period | Effective reproduction number for the index case | Symptomatic vs asymptomatic emissions | Ratio of aerosol vs. droplet emissions | Minimum close interaction time in cabins | Quarantine infection control efficiency | URT/LRT infectious doses |
| No. Scenarios | 10 | 5 | 6 | 2 | 3 | 2 | 2 | 3 |
| Range | 6 – 15 (days) | 1 – 5 (days) | 1 – 6 | 0.544<br>1.0 | 0.3:1<br>2.4:1<br>1:1 | 8 or 12 hours per day | Moderate<br>High | 1:1<br>10:1<br>100:1 |

## Results

A total of 132 model iterations met the acceptability criteria of R^2^ > 0.95 for daily cumulative cases and R^2^ > 0 for daily cases (0.6% of the total number of model iterations). The cumulative number of infected cases reported in various outlets was 765 cases; the average (±SD) cumulative number of modeled infected cases among iterations meeting acceptability criteria was 736 (±64) (Figure 1). A total of 611, 495, and 323 model scenarios achieved R^2^ > 0 for daily cases and R^2^> 0.8, 0.85, and 0.9 for daily cumulative cases, respectively.

**Figure 1.**
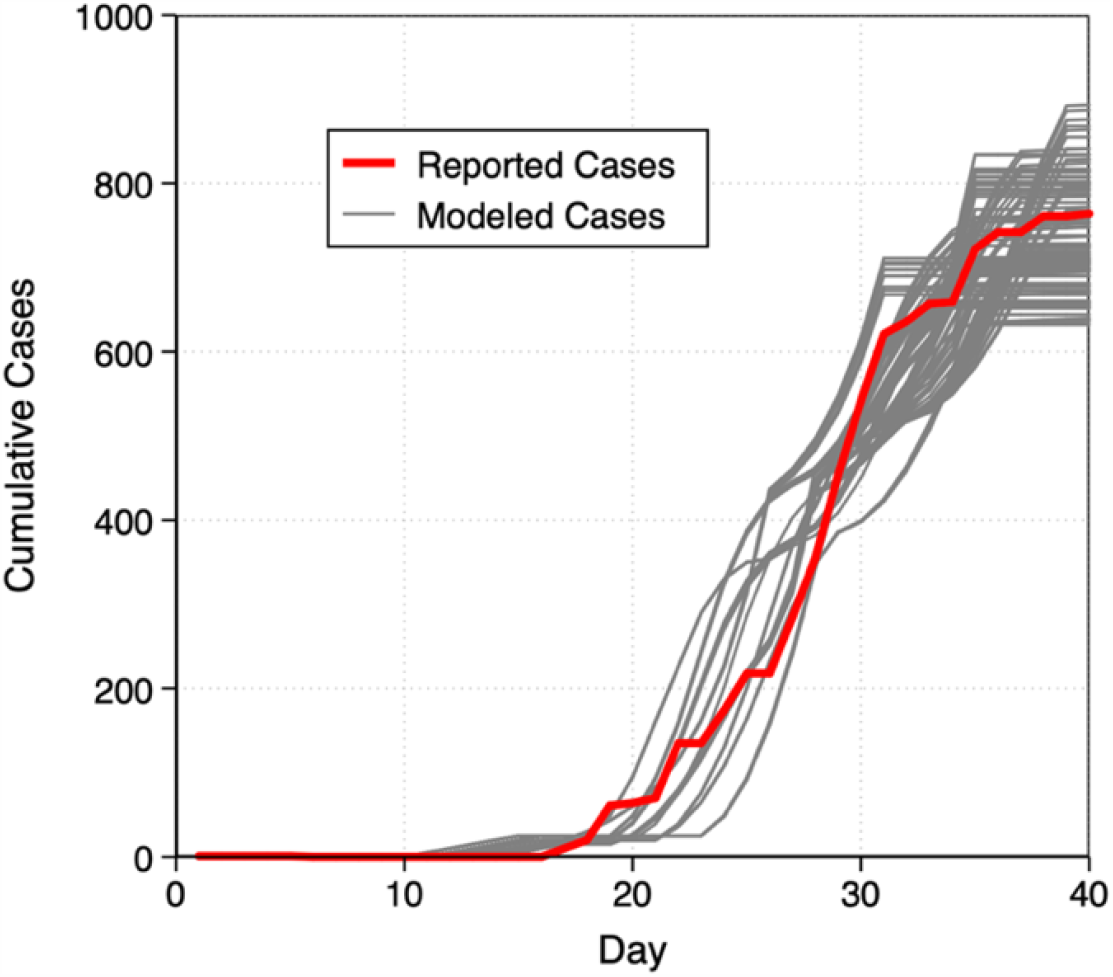
Reported (actual) and modeled (predicted) cumulative COVID-19 cases aboard the Diamond Princess Cruise Ship from January 20 – February 24, 2020. Modeled cases are from 132 model scenarios that met acceptable criteria (R^2^ >0.95 for cumulative daily cases and R^2^ > 0 for daily cases).

Table 2 shows the number of acceptable iterations that were associated with a specific assumption for each of the eight unknown or uncertain model input parameters, as well as the average R^2^ value for those iterations. Table 2 also shows the mean numerical estimate of each of these model input parameters, which demonstrates a ‘*best estimate*’ for each parameter using this approach.

**Table 2.** Distribution of acceptable model iterations (defined as R^2^ > 0.95 between reported and modeled daily cumulative case numbers and non-negative R^2^ for daily case numbers) that were associated with a specific assumption for eight unknown or uncertain model input parameters

| Model Inputs | Model Input Scenarios <sup>[a]</sup><br>Number of acceptable iterations (Average R<sup>2</sup>) |  |  |  |  | Best Estimates<br>(Mean ± SD) |
| --- | --- | --- | --- | --- | --- | --- |
| Effective incubation period | 10 days | 11 days | 12 days | 13 days | 14 days | 11.9 ± 1.3 |
|  | 25 (0.95) | 30 (0.97) | 31 (0.98) | 31 (0.97) | 15 (0.96) |  |
| Effective sub-clinical infectious period | 2 days | 3 days | 4 days | 5 days |  | 4.2 ± 1.1 |
|  | 14 (0.95) | 30 (0.97) | 1 (0.98) | 87 (0.97) |  |  |
| Asymptomatic / symptomatic emission scenarios <sup>[b]</sup> | A/S = 0.545 | A/S = 1.0 |  |  |  | 0.78 ± 0.23 |
|  | 64 (0.96) | 68 (0.98) |  |  |  |  |
| Emission rate scenarios (Aerosol/Droplet ratio) | A/D = 0.3 | A/D = 2.4 | A/D = 1.0 |  |  | A/D = 1.3 ± 0.9 <sup>[c]</sup> |
|  | 42 (0.97) | 50 (0.97) | 40 (0.97) |  |  |  |
| Minimum close interaction time in the cabins | 8 Hours | 6 Hours |  |  |  | 11.9 ± 4.0 |
|  | 68 (0.97) | 64 (0.97) |  |  |  |  |
| Effective reproduction number for the index case | R<sub>Eff</sub> = 2 | R<sub>Eff</sub> = 3 | R<sub>Eff</sub> = 4 | R<sub>Eff</sub> = 5 |  | 3.9 ± 0.9 |
|  | 11 (0.96) | 30 (0.97) | 53 (0.97) | 38 (0.97) |  |  |
| ID<sub>50</sub> URT / ID<sub>50</sub> LRT scenarios | ID = 1 | ID = 10 | ID = 100 |  |  | 47.1 ± 46.9 |
|  | 35 (0.97) | 39 (0.97) | 58 (0.97) |  |  |  |
| Infection control efficiency scenarios | Moderate | High |  |  |  | Moderate <sup>[d]</sup> |
|  | 70 (0.97) | 62 (0.97) |  |  |  |  |

[a] Scenarios with no acceptable model iterations were omitted from this table
[b] Asymptomatic refers to both asymptomatic and pre-symptomatic individuals
[c] The proportion of SARS-CoV-2 RNA copies emitted in form of inhalable aerosols (A) to large droplets (D)
[d] Non-numerical; thus, no number could be attributed as the mean value

Some estimates or assumptions for individual input parameters resulted in a larger proportion of successful model scenarios associated with that input compared to others (e.g., URT/LRT ID_50_ of 100:1, effective reproduction number of 4, effective sub-clinical infectious period of 5), which suggests that although these values may not be precise estimates or assumptions, they may be reasonably representative of the central tendencies of these parameters. Other parameters had similar numbers of successful model iterations associated with each assumed value, including effective incubation period, the ratio between asymptomatic (or pre-symptomatic) and symptomatic emission rates, aerosol/droplet emission ratios, minimum close interaction times in cabins, and infection control efficacy, which suggests that these parameters still have a high degree of uncertainty and/or may be less important for model sensitivity.

Figure 2 shows distributions of the estimated contributions of each transmission mode and viral source to the progress of COVID-19 aboard the ship over the entire duration that passengers remained aboard. Among the model scenarios meeting acceptability criteria, median (mean) estimates of the contributions of short-range (i.e., droplets and aerosols within close-range), long-range (i.e., aerosols outside of close-range contact), and fomite transmission modes to infected cases aboard the ship were 36% (35%), 41% (35%), and 21% (30%), respectively (Figure 2a). The estimated contribution of short-range (droplet + aerosol) transmission did not exceed 44% in any of the model scenarios that met acceptability criteria, while individual model scenarios exceeded 61% and 73% for long-range aerosol and fomite transmission, respectively. Conversely, the estimated contribution of short-range (droplet + aerosol) transmission was never lower than 22% for a single model scenario, while the estimated contributions of both long-range aerosol and fomite transmission were as low as 3% each, suggesting that the model framework yields a higher uncertainty in the contribution of short-range transmission than both long-range and fomite transmission. However, the central tendency of the most successful model iterations suggests that long-range aerosol and short-range aerosol + droplet transmission represented similar contributions to infection cases aboard the cruise ship, on average, with fomites likely contribution a smaller amount.

**Figure 2.**
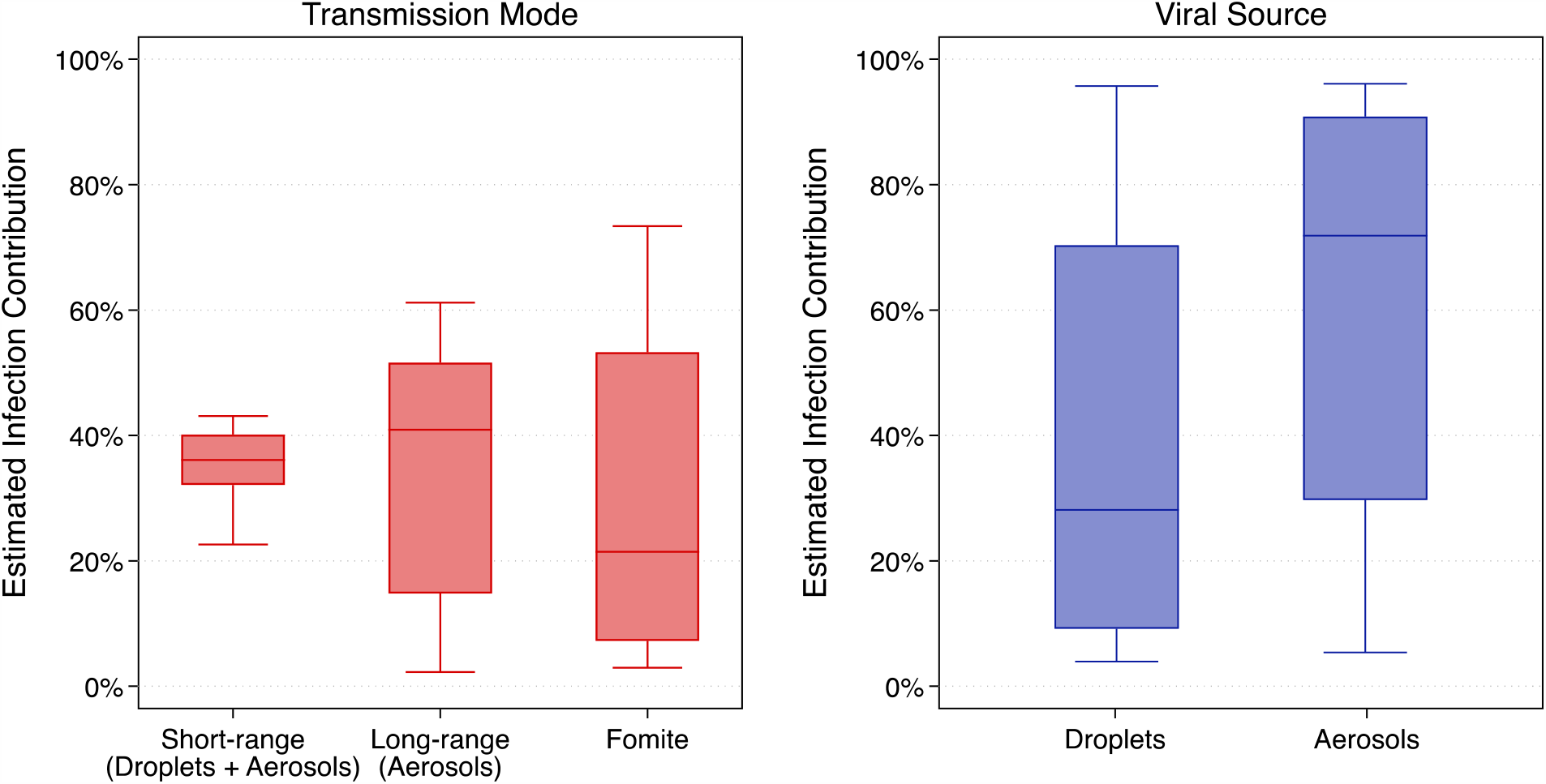
Estimates of the contributions of transmission modes and viral sources to infected cases aboard the Diamond Princess Cruise Ship over the entirety of the simulation period

Median (mean) estimates of the contributions of larger droplets (which includes only short-range and fomite transmission in the model framework) and smaller aerosols (which includes all possible modes of transmission) were 28% (41%) and 72% (59%), respectively (Figure 2b). Differences between droplet and aerosol transmission were significant (Mann-Whitney U-test p<0.0001). Individual model scenarios resulted in at least one scenario in which only one viral source dominated the other (up to 96% for each mode), but the central tendencies again suggest that smaller aerosols contributed to a greater proportion of infected cases aboard the cruise ship, on average, across all time periods (i.e., both before and after passenger quarantine).

Next, we analyzed the model results for periods before and after passenger quarantine started. Analyzing only the 132 model iterations that met acceptability criteria, the average (±SD) estimate of the proportion of cases that were transmitted prior to and after the passenger quarantine period was 58% (±5%) and 42% (±5%), respectively (Figure 3a). The average (±SD) estimate of the effective reproduction number before and after the quarantine period was 3.8 (±0.9) and 0.1 (±0.2), respectively (Figure 3b).

**Figure 3.**
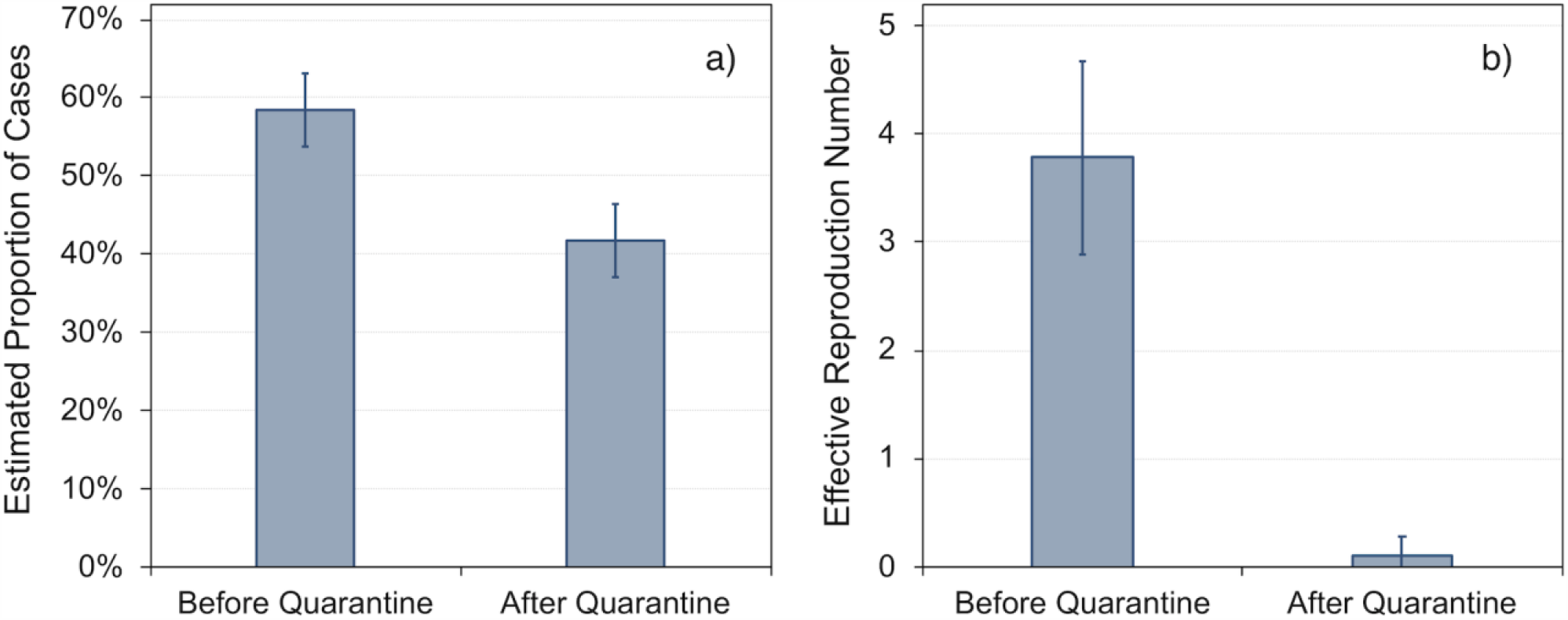
Mean (SD) estimates of (a) the proportion of cases and (b) the effective reproduction number before and after passenger quarantine

Estimates of the contributions of the specific transmission modes considered herein varied between the time periods before and after the passenger quarantine was in place (Figure 4). Prior to the passenger quarantine period, when passengers were free to move about both cabin and public areas, median (mean) estimates of the contribution of long-range, fomite, and short-range transmission were 42% (34%), 37% (46%), and 22% (19%), respectively, suggesting that close-contact transmission contributed the least to overall transmission, while long-range aerosol and fomite transmission were likely similar in magnitude. Conversely, after the quarantine period began and passengers primarily remained in their cabins, the median (mean) estimates of the contribution of long-range, fomite, and short-range transmission were 39% (36%), 58% (59%), and 0.5% (6%), respectively, suggesting that close-contact transmission (via both droplets and aerosols) dominated during this time period, as expected. Before the quarantine, only the differences between short- and long-range transmission (Mann-Whitney U-test p<0.0001) and between long-range and fomite transmission (Mann-Whitney U-test p=0.0004) were significant. After the quarantine, all transmission mode comparisons were significant (p<0.0001).

**Figure 4.**
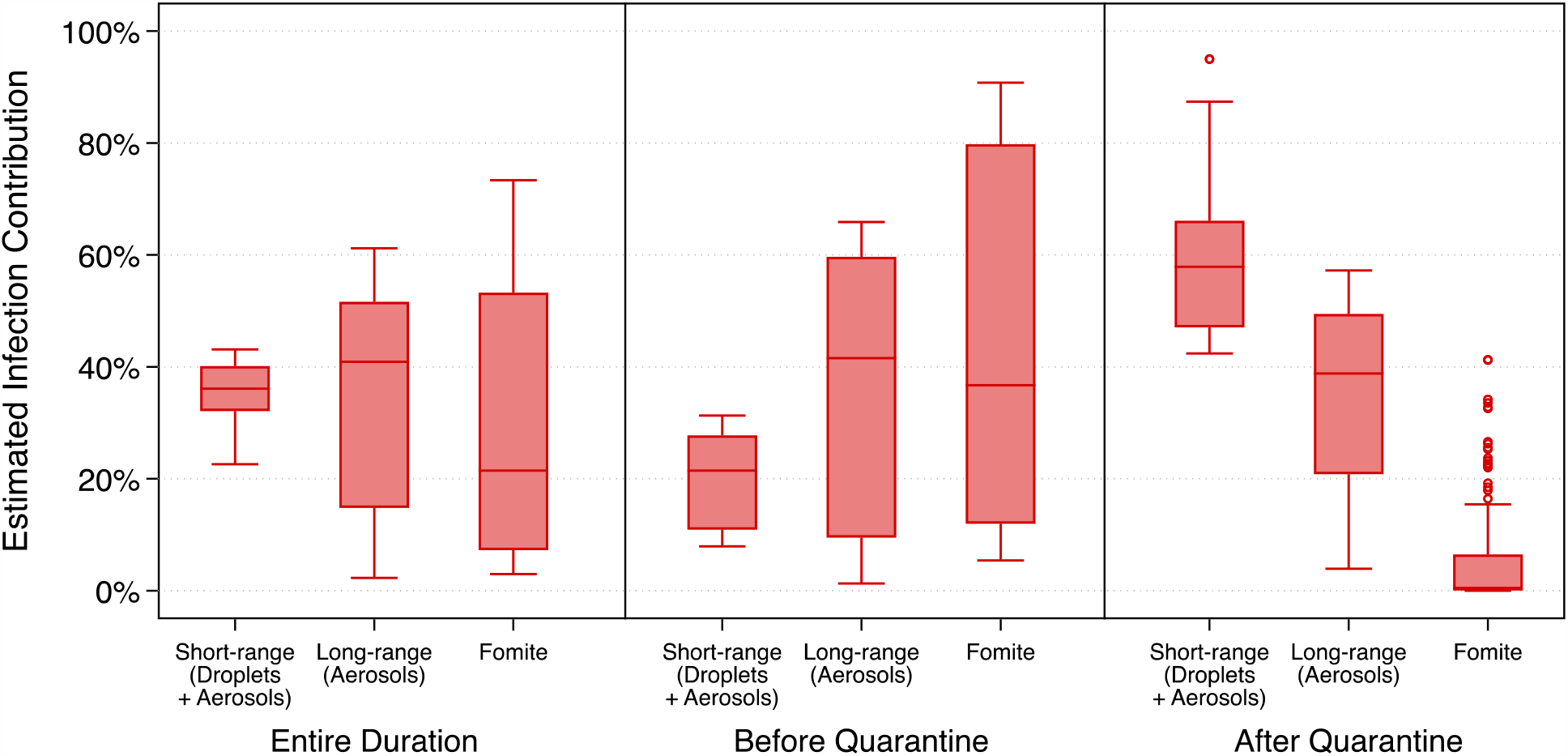
Estimates of the contribution of multiple transmission modes to infected cases aboard the Diamond Princess Cruise Ship over the entirety of the simulation period as well as before and after quarantine measures

Estimates of the contributions of the different viral sources considered herein (i.e., droplets vs. aerosols) also varied between the time periods before and after the passenger quarantine was in place (Figure 5). Median (mean) estimates of the contribution of droplets and aerosols prior to the passenger quarantine were 40% (50%) and 60% (50%) (p=0.32), respectively, suggesting that both larger respiratory droplets and smaller respiratory aerosols contributed approximately equally to infected cases aboard the ship during this time period. Conversely, median (mean) estimates of the contribution of droplets and aerosols after the passenger quarantine began were 15% (27%) and 85% (73%) (p<0.0001), respectively, suggesting that even though short-range transmission likely dominated during this period (Figure 4), smaller aerosol transmission likely accounted for the vast majority of infected cases post-quarantine, rather than larger droplets.

**Figure 5.**
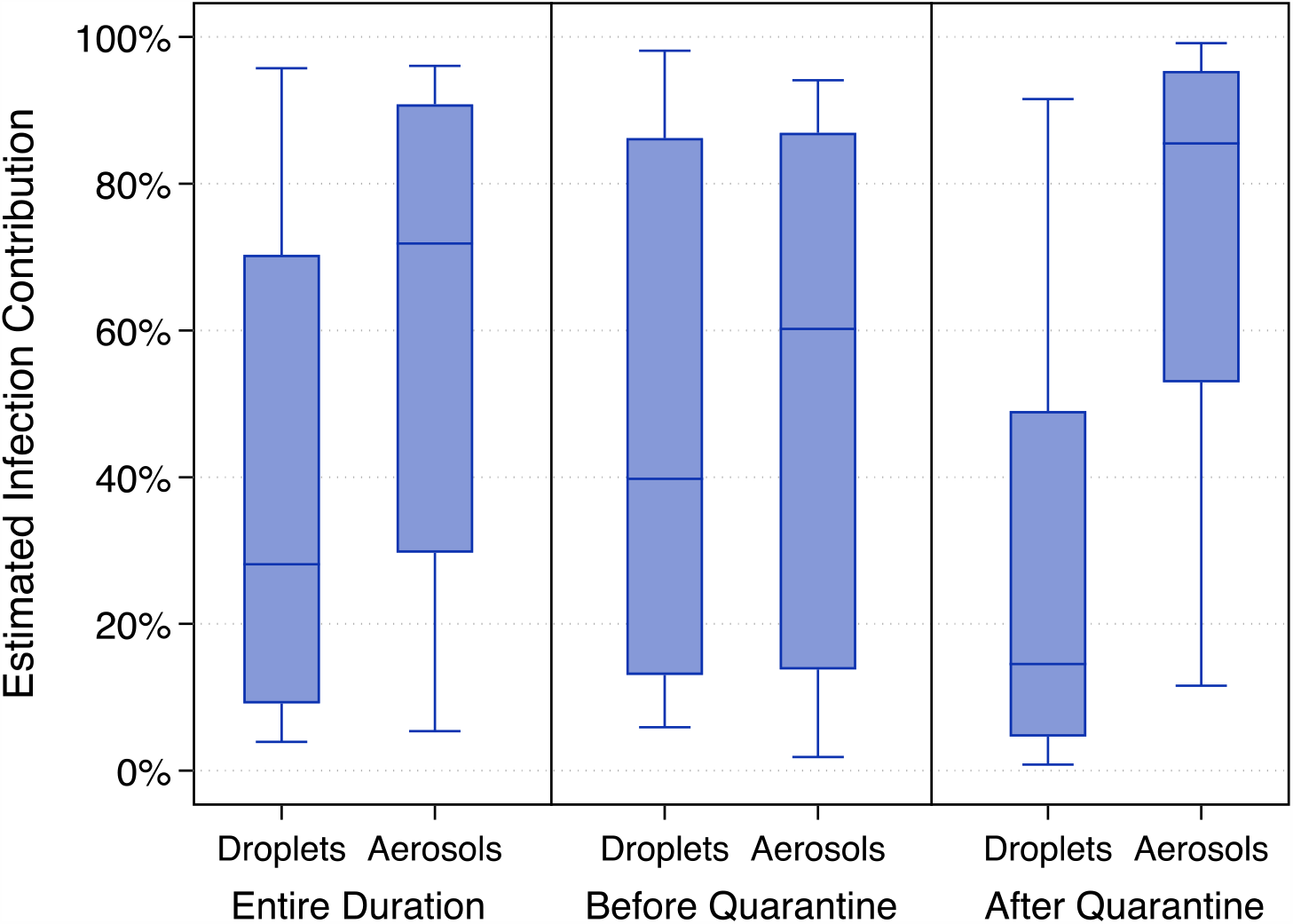
Estimates of the contribution of droplets and aerosols to infected cases aboard the Diamond Princess Cruise Ship over the entirety of the simulation period as well as before and after quarantine measures

## Discussion

Although there is high uncertainty around numerous model parameters, the model approach is designed to identify the most likely values of several unknown or uncertain parameters by analyzing only those model results that met acceptability criteria, and thereby providing insight into the likely importance of the various modes of transmission included in the framework. Results show that the long-range transmission of aerosols containing SARS-CoV-2 was most likely the dominant mode of COVID-19 transmission aboard the ship even with a very high ventilation rate (9-12 air changes per hour) and no recirculated air. The long-range and short-range transmission routes had similar contributions to the total number of infected cases. However, aerosol transmission across both short- and long-range distances accounted for >70% of disease transmission overall, which is contrary to the prevailing positions on how COVID-19 is spread.

Although cruise ships represent unique built environments with high ventilation rates and no air recirculation, these findings underscore the importance of implementing public health measures that target the control of inhalation of small aerosols in addition to ongoing measures targeting control of large droplet and fomite transmission. Moreover, our ‘*best estimates’* of the model parameters may be reasonably representative of the central tendencies of these parameters, particularly for estimates or assumptions of individual input parameters resulted in a larger proportion of successful model scenarios associated with that input compared to others as shown in Table 2.

We also conducted sensitivity analyses on the model results, described in detail in the SI (Section 4). Briefly, our sensitivity analyses demonstrate that: (i) aerosol transmission alone provides the strongest association between measured and reported cases in a mode elimination analysis (SI Section 4.2), (ii) primary epidemiological inputs among acceptable iterations most commonly clustered around effective sub-clinical infection periods of 5 days (with some 2-3 days) and effective incubation periods of 11-13 days (SI Section 4.3); (iii) the ratio between infectious dose of URT and LRT is a critical factor in the model and remains to be better understood from clinical investigations (SI Section 4.4); and (iv) the ratio for aerosol-to-droplet emissions remains an uncertain parameter, but has less influence on the results than the URT/LRT ID_50_ assumptions (SI Section 4.5).

There are several limitations to this modeling approach. For one, there is considerable uncertainty in our model inputs, as numerous estimates, assumptions, and implications were made because of a lack of available information, especially related to COVID-19 epidemic and mechanistic transmission characteristics, the interactions among individuals onboard the ship, and the effectiveness of infection control strategies adopted during the quarantine period. Some of these assumptions could have a significant impact on the results. For example, while the average contribution of fomite transmission among acceptable model iterations was estimated to be lower than other the other two pathways, under some specific assumptions (e.g., ID_50,URT_/ID_50,LRT_ = 1, see SI Section 4.4) or transmission periods (e.g., before passenger quarantine started), fomite transmission could have been the dominant transmission mode. Second, the model approach assumes constant and/or average values for numerous inputs in a given model iteration (e.g., every passenger was assumed to have the same probabilities of close-range contact with others and every infected individual was assumed to have the same emission rates of droplets in aerosols). By not considering variability in these parameters, we cannot directly account for “super-spreaders” and any underlying biological, physical, or behavioral differences in those individuals. Instead, the model framework produces average and uniform outcomes, which remains a limitation. Third, we relied on a conventional discrete size cut-off to define aerosols and droplets (i.e., 10 µm); however, respiratory droplets and aerosols actually exist on a continuum of particle sizes influenced by inertia, gravitational settling, and evaporation. We also did not consider the impacts of potentially influential characteristics such as temperature, humidity, sunlight, or not-well-mixed conditions in the control volumes considered herein. As more information becomes available, the model framework should continue to be tested and applied to other built environment transmission case studies.

## Contributors

The model development was led by PA, who came up with the original idea and advanced the framework with help from BRS, ZK, JGCL, and JGA. PA, ZK, and BRS contributed to processing the model and analyzing the outcomes. PA, BRS, and JGA contributed to writing the manuscript. PA, BRS, and ZK generated the figures and tables. All authors interpreted the findings and approved the final version for publication.

## Data Availability

All data used herein will be available, upon request, from the corresponding authors.

